# Formative Evaluation to Inform Recruitment and Engagement Strategies for emergency activation of blood donors in Nimba County, Liberia

**DOI:** 10.64898/2026.10.26.26366440

**Authors:** L. Ridge, L. Barnes, J.R. Lori, S. Kennedy, R. E. Ware, O. Subah, G. C. Zeantoe

**Affiliations:** University of Cincinnati; Linda S. Barnes Consulting; University of Michigan School of Nursing; UL-PIRE Center, University of Liberia; Cincinnati Children's Hospital and Medical Center; Academy of Public Health, West African Institute of Public Health; United Methodist University Winifred J. Harley School of Nursing

## Abstract

While donations from voluntary non-remunerated donors are preferred, approximately 14% of blood donations globally come from family replacement and paid donors. In low- and middle-income countries, the proportion is higher, averaging 36.6% in low-income countries. However, much less is known about family replacement and paid donors relative to voluntary non-remunerated blood donors. It is known that family replacement and paid donors are often activated in response to an emergency in areas where blood is scarce and whole blood is the standard for transfusion. Thus, they are part of de facto civilian walking blood banks. Interest in civilian walking blood banks is increasing, but many operational best practices have not been established. The objective of this study was to better understand the motivations and donation experiences of family replacement and paid donors, as well as their attitudes toward possible donor-facing walking blood bank practices. Semi-structured interviews were conducted in Nimba County, Liberia, in May 2026 with participants who had donated blood in the last twelve months. Recruitment was conducted through radio announcements and snowball sampling. Transcripts were analyzed via directed content analysis using the Exploration, Preparation, Implementation, and Sustainment framework. Twelve donors, 11 men and one woman, participated. Many reported making donations both as family replacement and as paid donors. Participants reported altruism, positive past donation experiences, and a desire to help their families as donation facilitators. They reported distance, concern about pre-transfusion testing, and expenses incurred as donors as barriers. While they reported widespread concern about the effects of blood donation on the donor’s health as a barrier to establishing a more formal walking blood bank approach, they typically believed outreach at community institutions would be an effective way to recruit new donors. These findings suggest that continued engagement among established donors in addition to recruitment of new donors is likely to increase the blood supply, although implementation questions around the best approaches to donor compensation and ensuring potential donors are paired with the closest facility remain.

## Objectives

A plentiful global blood supply is key to the ability to perform lifesaving transfusions (Asamoah-Akuoko et al., 2017; World Health Organization [WHO], 2026). However, not all areas collect the quantities of blood needed to support consistent transfusion access (Roberts et al., 2019; WHO, 2026). While the WHO recommends that all blood is obtained by a centralized blood collection system that relies on voluntary unpaid donors, about 13.7% of blood donations globally come from either family replacement or paid donors who are typically “activated” during an emergency to provide blood for a specific patient (WHO, 2026). The proportion of donations from family replacement or paid donors increases as countries’ incomes decrease; while in high-income countries 98.4% of donations are from voluntary unpaid donors, in low-income countries it is only 63.4% (WHO 2026). Despite their importance to low-income health systems, studies on family replacement and paid donors typically focus their rates of transfusion-transmitted infections relative to those of voluntary unpaid donors (Fernandes et al., 2010; Gadji et al., 2024). Meanwhile, studies on donor recruitment strategies emphasize centralized efforts to recruit donors in the context of high rates of voluntary unpaid donations (Schröder et al, 2025, Appiah et al., 2025).

Due in part to low rates of blood donation, some areas are simply unable to meet blood need. Areas that cannot meet 75% or more of their blood demand are known as blood deserts (Raykar et al., 2024). While the WHO recommends that all blood is obtained by a centralized blood collection system that relies on voluntary unpaid donors, in blood deserts other strategies merit exploration: drone delivery of blood, intraoperative autotransfusion, and civilian walking blood banks (Raykar et al., 2024).

Of these, civilian walking blood banks are likely the most aligned with current blood collection procedures in low-income countries. While implementation varies, civilian walking blood banks are characterized by three key practices: emergency activation, the use of rapid diagnostic testing, and the transfusion of whole blood (Raykar et al., 2024). One survey found that 77% of facilities in low- and middle-income countries rely on blood they have collected themselves to address acute patient needs, with 32% doing so exclusively (Barnes et al., 2022). According to the WHO (2024), 33 of 139 countries report that at least a percentage, and sometimes all, blood donations are tested using rapid diagnostic tests. The WHO (2026) reports that while 90% of donations globally were processed into components (e.g., platelets, plasma), this fell to 81% in lower-middle income countries and 52% in low-income countries. Thus, the research priority of developing implementable operational protocols and evidence-based best practices for civilian walking blood banks (Kumar et al., 2026; Raykar et al., 2024) has the potential to benefit many countries globally, including the United States (Brigmon et al., 2024; Kaada et al, 2019; Raykar et al., 2025).

Liberia, a country of 5.5 million people in West Africa, fits these patterns. Over 95% of blood collected in the country is collected at hospital-based blood banks (National Blood Safety Program (NBSP) Policy, 2023a). At the facility level, rapid diagnostic tests are the only tests available for transfusion-transmitted infections (NBSP Policy, 2023a). In both 2018 and 2023, zero whole blood donations in Liberia were processed into components (WHO, 2026). As a result of health system constraints, including very low rates of voluntary unpaid blood donation, fewer than 5% of Liberians live within 90 minutes of a transfusion-capable facility (Ridge et al., 2026), meeting the criterion for a blood desert. Thus, blood donor recruitment and retention are major national health priorities (NBSP Strategic Plan, 2023b). The objective of this study was to explore the experiences and motivations of family replacement and paid donors, and potential best practices for supporting civilian walking blood banks, in Liberia.

## Methods

This exploratory qualitative study used semi-structured individual interviews to investigate the donation experiences, motivations and deterrents to blood donation, and attitudes toward potential civilian walking blood bank-associated procedures among established blood donors in Liberia.

### Sample

Paid and family replacement donors were recruited through radio announcements and snowball sampling in Nimba County, Liberia. Nimba County is a blood desert where between 0.03 and 0.26% of residents live within 90 minutes of a transfusion-capable facility. Participants must have donated given blood within the last twelve months and be of 18 years of age or older.

### Instrument

The interview guide was developed by three researchers, two of whom had experience with data collection in Liberia and one of whom was an experienced Liberian researcher. The interview guide asked about recent donation experience, motivations and deterrents to giving blood, perceptions of the Liberian blood donation system, and opinions about walking blood bank-associated procedures such as recruitment.

### Data Collection

Data collection took place in May 2026. Informed consent was obtained prior to the interview. All interviews were conducted by both a researcher with experience in Liberia and the experienced Liberian researcher. Interviews were conducted in private offices at a local nursing school and the participating facilities. Interviews were tape recorded with the participant’s permission; if the participant declined to be recorded, handwritten notes were made. Recordings were sent via secure portal to a transcription service with experience in transcribing Liberian English, the Liberian variety of International English. Interviews ended when saturation, defined as when new examples of barriers and facilitators were no longer being generated, was reached (Saunders et al., 2018).

### Analysis

Transcripts were analyzed using directed content analysis, guided by the definitions of the Exploration, Preparation, Implementation, and Sustainment (EPIS) framework refined via a systematic review (Moullin et al., 2019). EPIS was chosen because of its inclusion of four ecological layers (outer context, bridging factors, innovation factors, and inner context) that, together, provided a comprehensive assessment. Specific EPIS factors were not included in the interview guide.

For the first round of coding, *Researcher One* read each transcript prior to coding for familiarity, then completed directed content analysis layer by layer. Codes were entered into comprehensive barrier and facilitator tables immediately after analysis. A second round of coding was conducted on *Author’s Institution’s Claude*, following best practices such as: explaining the study and going over the coding schema with the AI prior to any coding, ensuring transcripts were de-identified prior to uploading, uploading transcripts one at a time, and verifying all codes and quotations identified by AI using interview transcripts after coding is complete. *Author’s Institution’s Claude*, like *Researcher One*, generated two tables. All four tables were sent to *Researcher Tw*o for reconciliation.

This researcher was blinded to which were generated by a *Researcher One* and which by AI. *Researcher Two* then generated two comprehensive tables, one for barriers and one for facilitators.

### *Ethic*s

This study was approved by the University of Liberia’s IRB (Protocol# 2025-09-477). The author’s institution executed a reliance agreement with the University of Liberia.

## Results

Twelve donors participated, including 11 men and one woman. Interviews ranged from 12 to 24 minutes and averaged 17 minutes 46 seconds. Interviewees were student nurses (6), service workers (3), retired (1), or didn’t say (2). The categories of family replacement and paid donors were overlapping, as multiple donors reported both behaviors. All participants had donated when asked to do so for a specific patient emergency, as opposed to donating at a blood drive or blood bank. Many participants were repeat donors.

Some donors reported emergency activation despite not being the patient’s family member or having any prior donation history; rather, they had been recruited by hospital workers while on facility grounds for family visitation, work, or clinical rotations. These donors reported making the donation without requesting compensation. Donors had donated several different health facilities, including some outside of Nimba County.

### Facilitators

*Inner Context*. Donor-identified facilitators of the civilian walking blood bank approach were driven by the Inner Context (Table 1). In the individual context, the motivation to save lives was expressed by more donors than any other finding. Positive prior donation experiences, often related to low anxiety about donating, or a sense of personal accomplishment, also facilitated donations. The desire to help family was also prominent theme, though less common, and less straightforwardly a facilitator, since it was sometimes coupled with reluctance to donate to a stranger. Some donors also expressed a belief that donating was beneficial for their health, either as a direct result of the blood loss or as a result of obtaining free testing for transfusion-transmitted infections. Last, a sense of obligation among donors who were aware they had widely compatible blood types was also mentioned by multiple participants as important motivators. Donors often reported recruiting other people to donate, some of whom went on to become repeat donors.

**Table 1.**
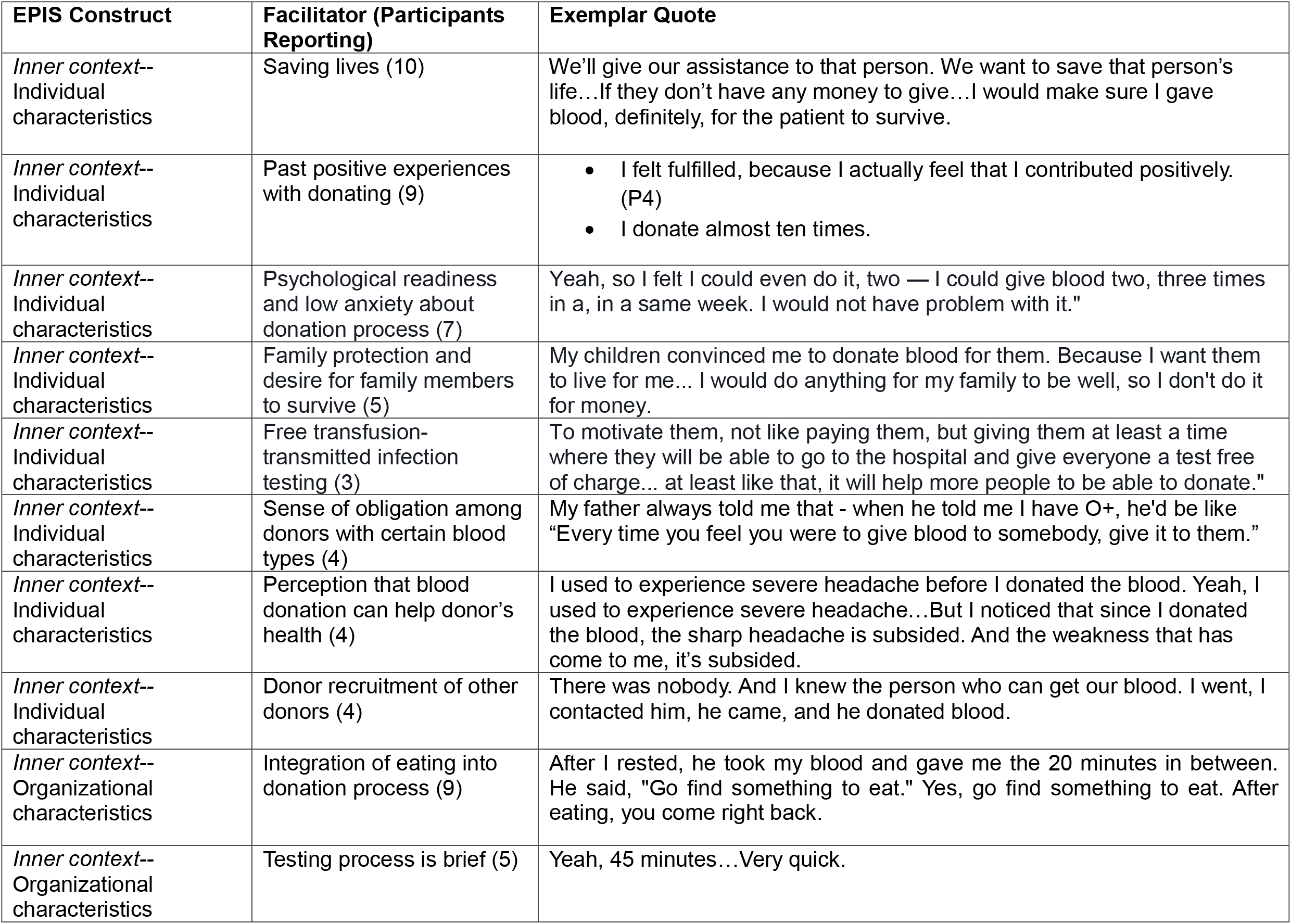
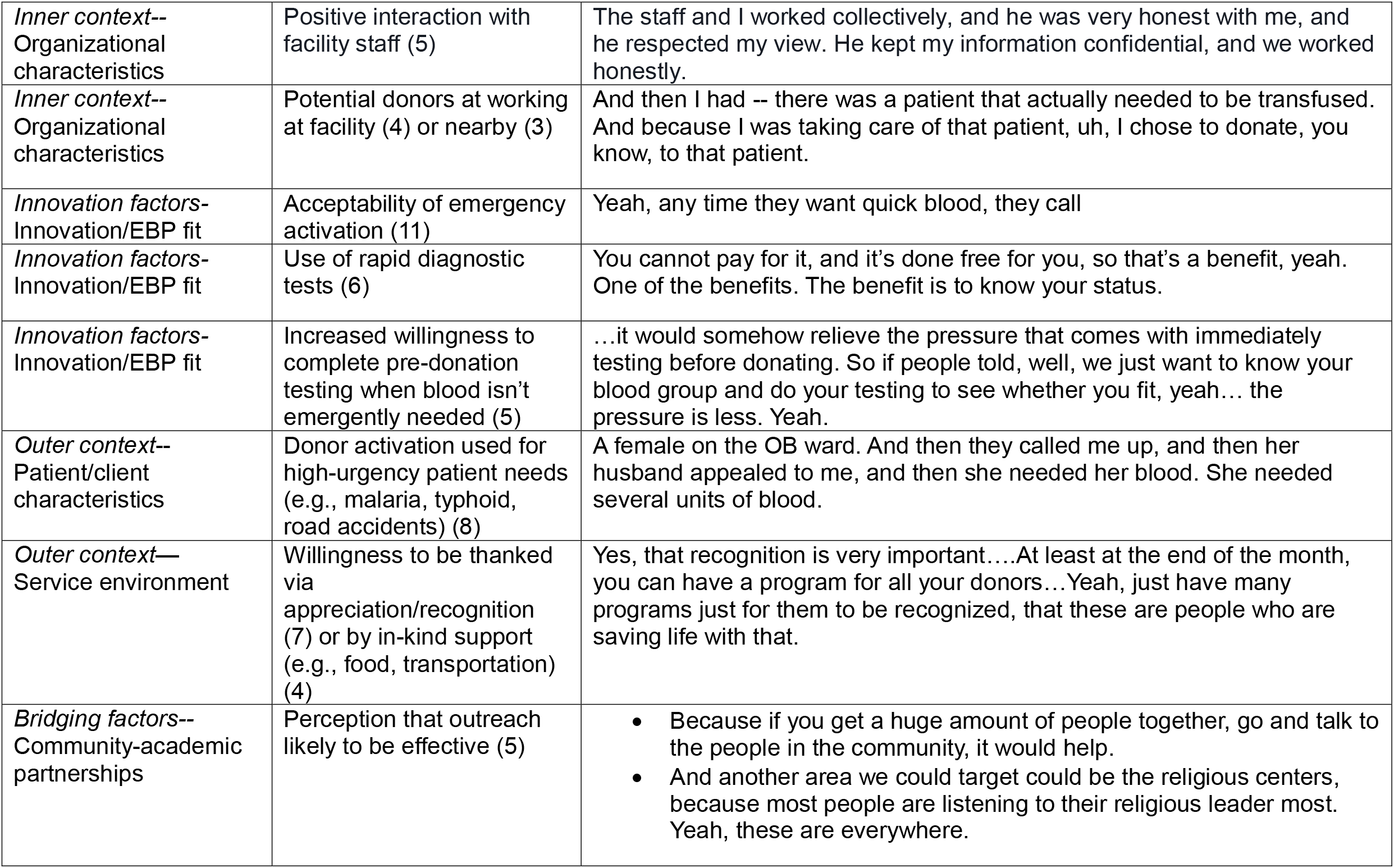
Facilitators of Blood Donation.

A few facilitators related to organizational characteristics. Donors frequently expressed appreciation that eating was integrated into the donation process, saying laboratory staff urged them to go eat between testing and the actual donation. Often the recommended was tomatoes and eggs. Other positive interactions with staff were mentioned, particularly during either the testing phase or the post-donation monitoring phase.

Donors also found the testing process acceptably brief. Last, several donors reported that proximity was a facilitator for them, as they were either recruited to donate because they were at the facility when the donation was needed (e.g., working, visiting an ill relative), or that they lived nearby.

#### Innovation Factors

The Innovation factor of “emergency activation” was highly acceptable among donors. One donor even expressed gratitude that he lived near the hospital and could be notified quickly in an emergency. Free rapid diagnostic testing was also mentioned as a facilitator.

One further innovation factor that was seen as a potential facilitator, although participants could not speak from personal experience, was that naïve donors might be more willing to complete testing for transfusion-transmitted infections when the donation was not needed (i.e., as part of a screening process to join a registry of potential donors).

#### Outer Context

The primary outer context facilitator was the use of emergency activation on behalf of patients with common, severe ailments, such as malaria (particularly with children), road accident trauma, and intrapartum blood loss.

The secondary outer context facilitator was that donors largely believed appreciation or recognition would be adequate compensation for many people. A few participants described personal thank-yous they had received from the blood recipient or their family as being particularly meaningful, while others thought more public recognition in the form of a displayable certificate or brief ceremony would be valuable. Donors also felt that often potential donors would be willing to accept compensation that only offset costs associated with the donation, like transportation and food.

#### Bridging factors

Promising bridging factors were the widespread belief that outreach, either at educational or religious institutions, or via the radio would effectively recruit more donors. One donor noted that outreach had been conducted in the past by the Government of Liberia and that it had generated a lot of interest at the time.

### Barriers

#### Outer Context

Barriers were primarily driven by the outer context (Table 2). The most frequently identified barrier was the costs incurred by the donor when making the donation. Two widespread beliefs were identified as further barriers: that blood donation had the potential to harm the donor’s health, and that donated blood is often sold in facilities. While some donors stated that they had previously personally been concerned blood donation would harm their health, none were still concerned. No donors reported personally believing that blood is sold after donation, rather than being administered the intended recipient.

**Table 2.** Barriers to Blood Donation.

| **EPIS Construct** | **Barrier (Participants)** | **Exemplar Quote** |
| --- | --- | --- |
| *Outer Context-Funding* | Expenses associated with the donation (e.g., food, transportation) (8) | • I would tell the person calling, oh, I don't have transportation. If they say come, I will pay, get on the motorbike, come, I will pay.<br>• To my place? To come is 150. To go back, it's 150. |
| *Outer Context-Service environment* | Belief that blood is sold (3) | Blood is expensive, it is sold in the hospital |
| *Outer context – Service environment* | Widespread perception of blood donation as harmful to health (5) | [After you have donated once,] if you don't do it [again], the blood will get excess in you. So people get afraid on that - on that notion. People are afraid to donate. Say if you give blood, you - it will be a regular process. If you not give it, then it will have effect on you. |
| *Inner context – Individual characteristics* | Distance to facility (6) | Well, the challenges might be, uh, firstly, the location of these people, where they are currently, because you said they're on the list. They could be called any time. But they are going on with their normal activities. |
| *Inner context – Individual characteristics* | Family donation preference (6) | Oh, that -- I'm done donating blood. I've been saving the family... I only help my family. |
| *Inner context – Individual characteristics* | Concern that the donation will negatively affect their health (6), particularly at the first donation (3) | I firstly will have the people check my blood level at the time. Because I want to make sure that it's enough, that even if I share, it wouldn't cause harm to me. So that's initially the first concern I normally put forth. |
| *Inner context – Individual characteristics* | Donor preference to give in exchange for cash (3) | The money is [there too]. Sometimes 3,000 Liberian dollars, now 4,000. |
| *Inner context - Individual characteristics* | Blood diversion concerns (3) | And another - another issue would be people at times, if a family will donate blood, and then that blood will not be transferred to the patient. |
| *Inner context-Organizational characteristics* | Process is too long (2) | So now -- so now like three hours, or four hours. |
| *Innovation factors-Innovation/EBP fit* | Anxiety around pre-transfusion testing (6) | Uh, the first time, I think one that caused me a bit scared was the one that I had to -- testing for HIV...testing for HIV, it really brings fear, so it was the one that I was a bit afraid, you know, why it is. But I ended up doing it, so -- it was negative. |
| *Innovation factors-Innovation/EBP fit* | Concerns about confidentiality of pre-testing results (2) | When I came to give the blood, I was nervous. Yeah, I was nervous. I was like, I was like, if these guys are going to know that I'm positive HIV, I would |

**Table 2. Barriers to Blood Donation**
|  |  |  |
| --- | --- | --- |
|  |  | feel... |

#### Inner Context

The primary inner context barrier was the donor’s distance from the facility when blood was needed. Another barrier identified with the same frequency was donor preference to give only to members of their own family. One donor who had given many times to his own children expressed reluctance to give to anyone unrelated.

Personal concerns, typically in the past, that the donation itself would negatively impact their health were also mentioned by many donors. A few donors mentioned the concern that donation would lead to an overabundance of blood in their bodies, requiring future donations to maintain health. Donors were definite that they no longer held these beliefs, but acknowledged they were once meaningful barriers. Last, donors were candid that either they themselves or others would prefer to receive cash in excess of their expenses in exchange for their donation; 3000 Liberian dollars (about $18 USD) was a figure given by a few participants. The only organizational barrier identified was that some participants found the donation process too time-consuming.

#### Innovation Factors

The innovation factor of “rapid diagnostic testing” was seen by many donors as a source of anxiety. Many donors reported anxiety associated with the actual results, particularly regarding HIV, while others reported concerns about potential violations of confidentiality. Some donors mentioned that their anxiety around testing decreased after the first donation.

## Discussion

These findings shed light on the factors motivating and deterring important, understudied populations of blood donors: family replacement and paid donors. They also identify several key donor-facing issues to consider when formalizing the implementation of civilian blood banks. The finding that many donors reported both family replacement and paid donation categories was not surprising, but is not well documented in the literature. It suggests that for some portion of emergency-activated donors, blood donation may become a part of their identity, as has often been documented among voluntary non-remunerated donors in Australia, Europe, and the United States (Edwards et al., 2022; France et al., 2014; Veldhuizen et al. 2013).

The most frequently cited facilitator of donation was altruism, consistent with previous studies in sub-Saharan Africa (Asamoah-Akuoko et al., 2017). Studies on donor attitudes in high-income countries with centralized blood donation found large groups of donors in these studies report low-to moderate levels of altruism (Schröder et al, 2025). While comparisons between these countries and low-income countries with decentralized blood collection is difficult, it is notable that Schröder et al. (2025) also found that the most altruistic donors are not the least likely to lapse. This suggests high rates of self-reported altruism might be an argument for continued engagement efforts rather than a basis for expecting sustained donations.

Many donors also reported positive past experiences as key facilitators. Specific aspects of prior experiences were consistently highlighted by these participants, including rapport with healthcare workers, whose encouragement that the donor eat prior to donation was appreciated, and experiences of personal fulfillment. While Nguyen et al. (2008) documented the high rates of satisfaction with individual workers among blood donors, only overall satisfaction was investigated for association with repeat donations. The effects of negative physiological responses to donation (e.g., syncope, dizziness) on future donation behavior has also been examined (France et al., 2013), but these findings suggest that previous donation experiences beyond the physical are important to repeat donation.

The main barriers identified by this group were distance to the facility and expenses incurred by blood donation. Prior studies have established that longer distance to blood donation facilities reduces donor retention in the context of voluntary non-remunerated blood donation (Piersma et al., 2021). The nature of emergency activation is such that distance would be even more of a factor for these donors. Thus, future efforts to establish a civilian walking blood bank would be wise to take distance into account when recruiting new donors by, for example, ensuring they are assigned to the facility nearest where they live or work, as opposed to where they are recruited.

The issue of expenses has ramifications for implementation. While donors consistently expressed willingness to accept recognition or appreciation as their main form of compensation, and cash did not come up as a significant motivator, as it has in other studies (Asamoah-Akuoko et al., 2017), the desire that donation costs be offset was regularly reported. The main expenses described are transportation and food, for which direct provision, vouchers, or cash compensation are often characterized as “low-value” incentives (Graf et al., 2024). In a context where transportation costs were reported as 150 Liberian dollars, or about one United States dollar, this characterization seems apt, and even suggests that compensating blood donors for these expenses might be a cost-effective intervention for health issues requiring blood transfusions, such as post-partum hemorrhage and accidents, which are drivers of morbidity and mortality in Liberia (Smith et al., 2019; WHO, 2023). Ideally, compensation for these expenses would be calibrated to avoid fueling commercialization or even just the perception that blood is bought. However, it is possible to take these concerns too far: countries of all income levels offer cash or cash-adjacent incentives such as tax relief, raffles, and gift cards (Graf et al., 2024). Thus, caution must be used to avoid unintentionally holding low-income countries to higher standards than appropriate.

These data identify further implementation issues of establishing civilian walking blood banks in low-income settings where emergency activation is already routine. First, that donors largely believe that outreach at high schools, colleges, and religious institutions is likely to be an effective recruitment strategy with the potential to grow the pool of blood donors contacted during emergencies. This belief is consistent with other investigations of blood donors in sub-Saharan Africa (Asamoah-Akuoko et al., 2017). Second, donors indicate that that this outreach can include rapid diagnostic testing, which is seen as a benefit by a significant portion of donors, and is also seen as likely to be more acceptable to potential first-time donors when completed before the donation is actually needed. The approach of rapid diagnostic testing at the time of enrollment has the dual benefit of expanding access to HIV testing in Liberia, a major public health goal (United Nations Liberia, 2022), and enhancing the safety of the blood supply, which at this time relies on a single round of rapid diagnostic tests used at hospital-based blood banks (NBSP Policy, 2023a).

Barriers to formalizing a walking blood bank approach were seen as anxiety around pre-donation testing, both in terms of the results themselves and potential lapses in confidentiality, widespread concerns about the health impact of blood donation, and expenses incurred by the donor as part of the donation process. Anxiety around pre-donation testing, particularly regarding HIV, has been previously documented in Liberia (Zimbizi, 2017) and in other countries in sub-Saharan Africa (Asamoah-Akuoko et al., 2017), and evidence-based approaches to minimize this anxiety, including pre-test counseling and privacy protocols, are key to walking blood bank implementation. The findings in this study that anxiety seems to abate significantly after the first donation, and that donors are often happy to recruit other donors, suggest current donors may have a meaningful role to play. Widespread concerns about the health impact of blood donation have also been documented in Liberia (Zimbizi, 2017) and globally (Bednall & Bove, 2011) previously. Again, donors often reported a personal history of these concerns that abated after their first donation. The fact that two such significant barriers are lowered so meaningfully by donating, even once, suggests that efforts to increase the rate of blood donation that focus on established donors are at least as likely to be successful as efforts that focus on naïve donors.

### Limitations

It is possible that social desirability bias influenced participants’ answers, particularly regarding the role of financial incentives as a source of motivation. However, participants were candid about several less altruistic preferences, such as preferring to donate only to family. Additionally, our participants were overwhelmingly male. Liberia’s National Blood Safety Policy (2024) states blood donors in Liberia are 77% male, but our sample was more than 90% male. This may have limited our ability to identify issues primarily affecting female donors.

## Conclusion

This study explored the perceptions and motivations of family replacement and paid donors, a substantial group of blood donors who are understudied compared to their voluntary unpaid counterparts. Donors often had a history of donating both as family replacement and paid donors. Important motivations were altruism and positive past donation experiences. Important barriers were misconceptions about the risks of blood donation, fears regarding testing for transfusion-transmitted infections, and the preference to give only to one’s own family. The first two barriers were attenuated by the donation experience. Donor perceptions relevant to establishing evidence-based best practices for civilian walking blood banks in Liberia and elsewhere included high acceptability of emergency activation, widespread beliefs about the potential harms of blood donation, widespread concerns about pre-donation testing, and the belief in the efficacy of outreach in recruiting new donors. Since two of these barriers are lowered by experience with donating blood, whether outreach efforts should focus on new or established donors merits further investigation.

## Data Availability

All data produced in the present study are available upon reasonable request to the authors

## Notes

### Competing Interest Statement

The authors have declared no competing interest.

### Author Declarations

IRB of University of Liberia gave ethical approval for this work IRB of University of Cincinnati gave ethical approval for this work

### Funding

- University of Cincinnati

## References

Appiah, B., Asamoah-Akuoko, L., Ahmed, M. K., Tancred, T., Dei-Adomakoh, Y., Singh, G., Reilly, C., Olayemi, E., Delaney, M., France, C. R., Benneh-Akwasi Kuma, A., Yawson, A. E., Telke, S., Asiedu-Danso, M., Dei, E. N., Adongo, P., & Bates, I. (2025). Efficacy of communication interventions for promoting blood donation in low- and middle-income countries: A systematic review. Vox Sanguinis, 120(5), 432–446. 10.1111/vox.13799

Asamoah-Akuoko, L., Hassall, O. W., Bates, I., & Ullum, H. (2017). Blood donors’ perceptions motivators and deterrents in Sub-Saharan Africa - a scoping review of evidence. British Journal of Haematology, 177(6), 864–877. 10.1111/bjh.14588

Barnes, L. S., Stanley, J., Bloch, E. M., Delaney, M., Spitalnik, S. L., & Raykar, N. P. (2022). Status of hospital-based blood transfusion services in low-income and middle-income countries: A cross-sectional international survey. BMJ Open, 12(3), Article e055017. 10.1136/bmjopen-2021-055017

Bednall, T. C., & Bove, L. L. (2011). Donating blood: A meta-analytic review of self-reported motivators and deterrents. Transfusion Medicine Reviews, 25(4), 317–334. 10.1016/j.tmrv.2011.04.005

Brigmon, E. P., Cirone, J., Harrell, K., Cap, A. P., Gurney, J. M., & Spinella, P. C. (2024). Walking blood bank: A plan to ensure self-sufficiency in an era of blood shortage. Trauma Surgery & Acute Care Open, 9(Suppl 1), Article e001151. 10.1136/tsaco-2023-001151

Edwards, A. R., Masser, B. M., & Barlow, F. K. (2023). Psychological ownership and identity motives in blood donation. Vox Sanguinis, 118(8), 616–623. 10.1111/vox.13479

Fernandes, H., D’souza, P. F., & D’souza, P. M. (2010). Prevalence of transfusion transmitted infections in voluntary and replacement donors. Indian Journal of Hematology and Blood Transfusion, 26(3), 89–91. 10.1007/s12288-010-0044-0

France, C. R., France, J. L., Wissel, M. E., Ditto, B., Dickert, T., & Himawan, L. K. (2013). Donor anxiety, needle pain, and syncopal reactions combine to determine retention: A path analysis of two-year donor return data. Transfusion, 53(9), 1992–2000. 10.1111/trf.12069

France, C. R., Kowalsky, J. M., France, J. L., Himawan, L. K., Kessler, D. A., & Shaz, B. H. (2014). Blood donor identity survey. Transfusion, 54(8), 2098–2105. 10.1111/trf.12588

Gadji, M., Gueye, Y. B., Motto, D., & Diop, S. (2024). Seroprevalence of transfusion-transmissible infections among family replacement donors and voluntary non-remunerated blood donors during the COVID-19 pandemic in Sub Saharan Africa. Mediterranean Journal of Hematology and Infectious Diseases, 16(1), Article e2024008. 10.4084/MJHID.2024.008

Graf, C., Oteng-Attakora, K., Ferguson, E., Vassallo, R., & Merz, E.-M. (2024). Blood donor incentives across 63 countries: The BEST Collaborative study. Transfusion Medicine Reviews, 38(2), Article 150809. 10.1016/j.tmrv.2023.150809

Kaada, S. H., Apelseth, T. O., Hagen, K. G., Eliassen, H. S., Kristoffersen, E. K., & Hervig, T. A. (2019). How do I get an emergency civilian walking blood bank running? Transfusion, 59(S2), 1446–1452. 10.1111/trf.15184

Kumar, N., Mathew, F., Sawhney, R., Thompson, A. A., Makanga, C., Williams, W., Wesonga, C., Musa, A., Nyanamba, N. O., Lokuruka, K. K., Barmasai, H., Wakwabubi, M., Wangia, E., Opiyo, C. O., Esekon, E., Gilchrist, L., Puyana, J. C., Kumar, P., Delaney, M., … Barnes, L. (2026). Implementation and evaluation of a community walking blood bank for haemorrhagic shock when banked blood is unavailable: Protocol for a mixed-methods, effectiveness-implementation study - phase 2 of the Local Initiative for emergency blood (LIFE-Blood) study. BMJ Open, 16(7), Article e113702. 10.1136/bmjopen-2025-113702

Ministry of Health. (2023a). National blood safety policy 2024–2033. Republic of Liberia.

Ministry of Health. (2023b). National blood safety strategic plan 2024–2028. Republic of Liberia.

Moullin, J. C., Dickson, K. S., Stadnick, N. A., Rabin, B., & Aarons, G. A. (2019). Systematic review of the Exploration, Preparation, Implementation, Sustainment (EPIS) framework. Implementation Science, 14(1), Article 1. 10.1186/s13012-018-0842-6

Nguyen, D. D., DeVita, D. A., Hirschler, N. V., & Murphy, E. L. (2008). Blood donor satisfaction and intention of future donation. Transfusion, 48(4), 742–748. 10.1111/j.1537-2995.2007.01600.x

Piersma, T. W., Bekkers, R., de Kort, W., & Merz, E.-M. (2021). Altruism in blood donation: Out of sight out of mind? Closing donation centers influences blood donor lapse. Health & Place, 67, Article 102495. 10.1016/j.healthplace.2020.102495

Raykar, N., Chelagat, T., Barnes, L. S., Kennedy, S. S., Subah, M., & Delaney, M. (2025). The local initiative for emergency blood (LIFE-blood) study: A mixed-methods, single-center exploration of civilian walking blood bank need, feasibility, and safety in a low-resource blood desert. Transfusion, 65(12), 2293–2305. 10.1111/trf.18460

Raykar, N. P., Raguveer, V., Abdella, Y. E., Al-Riyami, A. Z., Bloch, E. M., Cap, A. P., Delaney, M., Spitalnik, S. L., & Blood DESERT Coalition. (2024). Innovative blood transfusion strategies to address global blood deserts: A consensus statement from the Blood Delivery via Emerging Strategies for Emergency Remote Transfusion (Blood DESERT) Coalition. The Lancet Global Health, 12(3), e522–e529. 10.1016/S2214-109X(23)00564-8

Ridge, L. J., Norrod, P., Barnes, L., Kennedy, S., & Subah, O. (2026). Identifying blood deserts in Liberia: A geospatial analysis. Transfusion. Advance online publication. 10.1111/trf.70292

Roberts, N., James, S., Delaney, M., & Fitzmaurice, C. (2019). The global need and availability of blood products: A modelling study. The Lancet Haematology, 6(12), e606–e615. 10.1016/S2352-3026(19)30200-5

Saunders, B., Sim, J., Kingstone, T., Baker, S., Waterfield, J., Bartlam, B., Burroughs, H., & Jinks, C. (2018). Saturation in qualitative research: Exploring its conceptualization and operationalization. Quality & Quantity, 52(4), 1893–1907. 10.1007/s11135-017-0574-8

Schröder, J. M., Ramondt, S., Spekman, M., & Merz, E. M. (2026). From unfulfilled to exceptionally motivated: Motivational types, change, and retention of blood and plasma donors. Psychology & Health, 41(9), 1289–1320. 10.1080/08870446.2025.2519113

Smith, J. M., Baawo, S. D., Subah, M., Sissoko, D., & Gibson, H. (2014). Advance distribution of misoprostol for prevention of postpartum hemorrhage at home births in two districts of Liberia. BMC Pregnancy and Childbirth, 14(1), Article 189. 10.1186/1471-2393-14-189

United Nations Liberia. (2022). Joint UN programme of support on AIDS 2022–2026 for Liberia. https://liberia.un.org/sites/default/files/2022-02/Joint%20UN%20Programme%20of%20Support%202022-2026%20_%20Final_%209Feb22.pdf

Veldhuizen, I. J. T. (2013). Blood donor profiling using donation patterns. ISBT Science Series, 8(1), 233–237. 10.1111/voxs.12017

World Health Organization. (2023). Global status report on road safety 2023. https://cdn.who.int/media/docs/default-source/country-profiles/road-safety/road-safety-2023-lbr.pdf?sfvrsn=60b635e7_3&download=true

World Health Organization. (2025). Global database on blood safety. https://www.who.int/teams/health-product-policy-and-standards/standards-and-specifications/blood-and-products-of-human-origin/quality-and-safety/global-database-on-blood-safety

World Health Organization. (2026). Global status report on blood safety and availability 2025.

Zimbizi, G. (2017). ACCEL Project: Informing blood donor mobilization strategies: Assessment on knowledge, attitudes, behaviour and practices of blood donors, potential donors and health care workers concerning blood donation in Liberia. ACCEL Project.

